# The stool microbiome in patients with psoriatic arthritis is altered but, unlike the skin microbiome, does not change following treatment: evidence for an underlying inflammatory drive from the intestine

**DOI:** 10.1101/2023.05.18.23289979

**Authors:** Madhura Castelino, Umer Z Ijaz, Mauro Tutino, Elizabeth MacDonald, George E. Fragoulis, Pauline Ho, Devesh Mewar, Gladston Chelliah, Andrew McBain, Catherine O’Neill, Stefan Siebert, Anne Barton, Simon Milling

## Abstract

**Objectives:** There is growing recognition that interactions between bacteria and the immune response may contribute to inflammation in psoriatic arthritis (PsA). To identify how the skin and stool microbiota may contribute to pathogenesis, we analysed the microbiota and immunophenotype of patients with PsA before and after therapy.

**Methods:** We examined the phenotype of PBMCs as well as skin and stool microbiota from 22 healthy volunteers, 7 PsA patients receiving methotrexate, and 23 PsA patients treated with biologic DMARDs. Samples for microbiome analysis were collected before therapy and after 3-6 months. First, we identified differences in the skin and stool microbiota between PsA patients and health volunteers. We then assessed changes in the microbiome occurring after treatment.

**Results:** As hypothesised, treatment responses were reflected by changes in both the skin microbiota and in the immunophenotype. However, in the stool samples, dysbiosis persisted after therapy. This dysbiosis was associated with changes in the peripheral blood immunophenotype. Correlation analysis enabled identification not only of specific microbial taxa (Clostridial species) that contributed to the persistent dysbiosis, but also of interactions between taxa and the immune response.

**Conclusions:** These analyses indicate that specific Clostridial species contribute to persistent gut dysbiosis in PsA, and their prevalence is associated with specific immunological changes that are not altered by treatment. Thus, an underlying inflammatory response in the intestine appears to contribute to the pathogenesis of PsA.

**Key Messages:** What is already known about this subject?

- Intestinal bacterial populations are altered in people with inflammatory diseases, including psoriatic arthritis (PsA).
- Interactions between bacteria and the immune system may contribute to the pathogenesis of inflammatory diseases.

What does this study add?

- While skin bacteria in patients with PsA revert towards normal after treatment, stool bacteria remain different from healthy individuals.
- Specific bacterial types are correlated with specific changes in the immune system in PsA.

How might this impact on clinical practice or future developments?

- Identification of the bacteria that drive persistent underlying changes in the immune response may enable these to be targeted.

## Introduction

Psoriatic arthritis (PsA) is a chronic inflammatory arthritis characterised by musculoskeletal and skin involvement. There is an increasing recognition that changes in the human microbiome may contribute to the pathogenesis of many chronic inflammatory conditions. The human microbiome, defined as micro-organisms, their genomes as well as the surrounding environmental conditions (1)can therefore be considered a potential modifiable factor in these conditions. There are several lines of evidence suggesting a potential role of microbial interactions in PsA. Scher et al reported gut dysbiosis (n=16) in recent onset, treatment-naïve PsA patients with reduction of taxa (*Akkermansia audiophilia* and *Ruminococcus*) that produce short-chain fatty acids compared to healthy controls(2). In addition, changes observed in the skin microbiota in psoriatic skin plaques(3), along with inflamed and abnormal skin barrier in psoriasis(4),may play a contributory factor in the pathogenetic mechanisms of inflammation and immune response in PsA.

Recent work has indicated that the microbiome may affect treatment response in rheumatoid arthritis (RA)(5) Crohn’s disease(6) and cancer immunotherapy(7). Furthermore, microbial composition has been reported to be altered in RA treated with conventional synthetic disease-modifying anti-rheumatic drugs (csDMARDs)(8) and biologic DMARDs (bDMARDs)(9).

We hypothesise that response to DMARD treatment in PsA is associated with changes in the skin and stool microbiome, which may be mediated by changes in the host immune response. The aims of this work were to investigate the relationship between the skin and stool microbiota and the host immune response in individuals with PsA, and to identify any changes that occur after initiation of treatment with methotrexate (MTX) or bDMARDs.

## Methods

### Study participants

Patients with PsA, fulfilling the CASPAR criteria(10) starting on MTX or first bDMARD as part of standard clinical care were recruited from four centres (3 in the North-West of England and one from Scotland) with approved study protocols (West of Scotland REC 4 14/WS/1035 North West 13/NW0068). Disease activity was assessed using PsARC before and after treatment. Age and gender matched healthy volunteers were recruited through the University of Manchester (MREC 99/8/84). Informed written consent was obtained from all participants.

Exclusion criteria for healthy volunteers included a history of antibiotic use within three months of sampling, probiotics use or known history of chronic inflammatory disease. PsA patients were excluded if they had received antibiotics within three months of the baseline visit. If antibiotics were used after the initial recruitment, the subsequent sampling was deferred for at least three months from completion of the antibiotic treatment.

Sample collection

PsA patients provided samples at two time points: before commencing and three to six months after starting treatment. Samples were collected from healthy volunteers on one occasion only. Skin swabs were obtained bilaterally from the extensor aspect of the upper and lower limbs and the scalp in the healthy volunteers and from lesional skin at the same 6 skin sites, where affected by psoriasis, in participants with PsA. Samples were collected using pre-moistened swabs and were frozen at -20°C within 1 hour of collection. Peripheral blood for immunophenotyping was collected at the same time as the skin samples, from all participants with PsA and the healthy volunteers. Stool samples were collected at home by the participants then posted to the University of Manchester in RNALater and subsequently frozen at -20°C.

### Sample processing

#### 16S rRNA gene sequencing

Stool and skin bacterial DNA was extracted and the V4 hypervariable region of the 16S rRNA gene was amplified and sequenced on MiSeq Benchtop sequencer as described previously(11). Fastq files were pre-processed using a bespoke amplicon analysis pipeline(12) and OTU assignment was performed using RDP classifier and SILVA database v123.

#### Immunophenotyping

Peripheral Blood Mononuclear Cells (PBMCs) were isolated within 3 hours of blood collection using 50mL Leucosep tubes and Histopaque (Sigma-Aldrich) and stored at -80°C in 20% DMSO Foetal Calf Serum (FCS; Invitrogen). During PBMC isolation, plasma was extracted and stored at -80°C. All samples were analysed using the same flow cytometry instrument.

#### Statistics

Detailed descriptions of all statistical methods are presented in the supplementary information. Statistical analyses were performed in R, using the vegan package for alpha and beta diversity analyses. Discriminant analyses, to identify microbial differences between groups, were performed using different algorithms for the stool and skin. For the stool samples, Sparse Projection to Latent Structure – Discriminant Analysis (sPLS-DA) from the R mixOmics package was used. For the skin, where samples were collected from three sites, the Multivariate Integration (MINT) algorithm was applied. Discriminant analyses to identify correlations between individual elements of the microbiome and immunophenotype were undertaken with DIABLO from the R mixOmics package. In figures displaying boxplots, pair-wise ANOVA was performed.

## Results

### Clinical characteristics of the participants

Thirty-two participants with PsA (seven commencing MTX and 25 commencing bDMARD) and 22 age-and sex-matched healthy volunteers were recruited and provided samples, however 2 participants from the bDMARD group were not included in the final analysis as one withdrew from the study and one had to defer starting the treatment due to other medical problems (Table 1). The bDMARDs were: adalimumab (7), etanercept (11) secukinumab (4) and cetrolizumab (1). At recruitment, participants with PsA had a higher median body mass index (BMI) than the healthy volunteers. There were more patients with a history of smoking (current or ex-smokers) among those commencing on bDMARDs. Disease duration and disease activity were lower in those commencing MTX than those starting bDMARDs (Table 1).

**Table 1.**
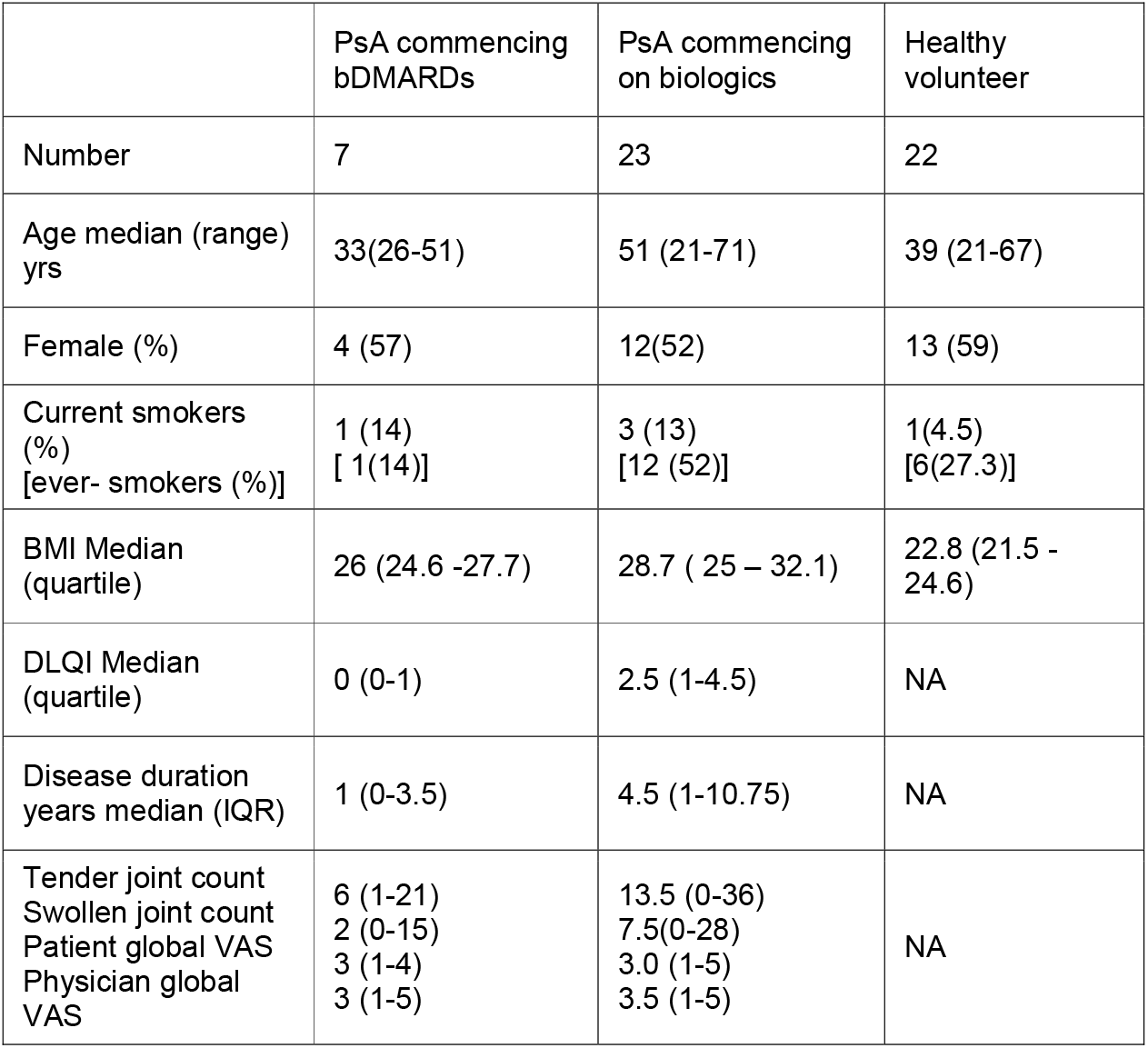
Baseline Demographics of study participants. PsA – Psoriatic Arthritis; BMI = Body Mass Index; DLQI = Dermatology Life Quality Index; IQR = Interquartile Range; VAS = visual analogue scale.

|  | PsA commencing bDMARDs | PsA commencing on biologics | Healthy volunteer |
| --- | --- | --- | --- |
| Number | 7 | 23 | 22 |
| Age median (range) yrs | 33(26-51) | 51 (21-71) | 39 (21-67) |
| Female (%) | 4 (57) | 12(52) | 13 (59) |
| Current smokers (%)<br>[ever- smokers (%)] | 1 (14)<br>[ 1(14)] | 3 (13)<br>[12 (52)] | 1(4.5)<br>[6(27.3)] |
| BMI Median (quartile) | 26 (24.6 -27.7) | 28.7 ( 25 – 32.1) | 22.8 (21.5 - 24.6) |
| DLQI Median (quartile) | 0 (0-1) | 2.5 (1-4.5) | NA |
| Disease duration years median (IQR) | 1 (0-3.5) | 4.5 (1-10.75) | NA |
| Tender joint count<br>Swollen joint count<br>Patient global VAS<br>Physician global VAS | 6 (1-21)<br>2 (0-15)<br>3 (1-4)<br>3 (1-5) | 13.5 (0-36)<br>7.5(0-28)<br>3.0 (1-5)<br>3.5 (1-5) | NA |

At the follow-up visit, 43% of the participants in the MTX and 74% of the participants in the bDMARD groups were classified as responders based on the improvement in their PsARC criteria.

### Alpha and beta diversity indices in skin and gut in PsA

At baseline, before treatment, no differences were observed in alpha diversity metrics (representing mean species diversity) in individuals with PsA (M1 and B1) compared to healthy volunteers (HV); neither the richness (species counts) nor Shannon Index (species proportional abundances) differed between subjects with PsA and healthy volunteers when microbial communities of the skin and stool were considered (Figure 1: A-D).

**Figure 1:**
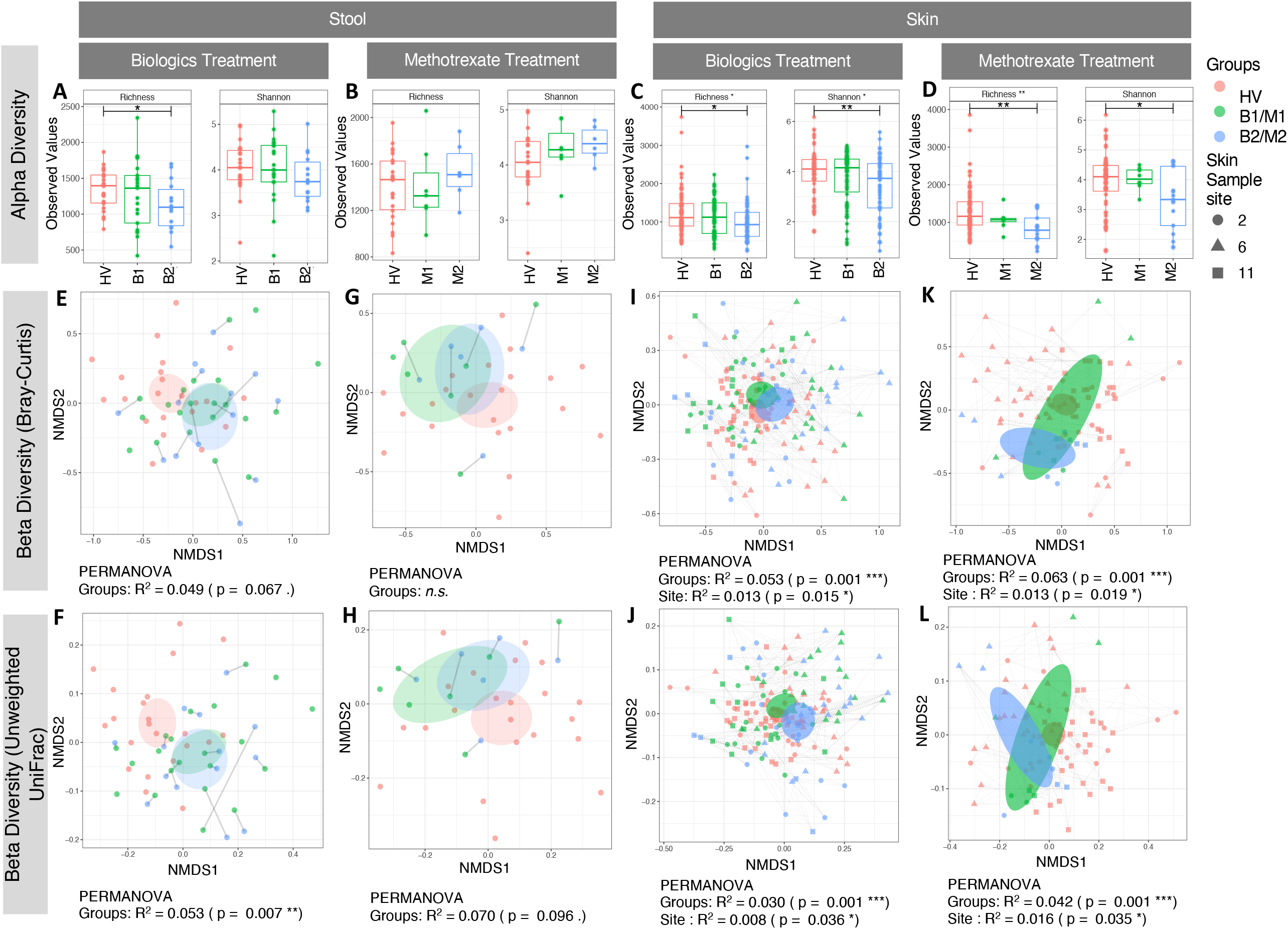
Microbial alpha diversity and beta diversity estimates. **(A)** Stool microbiome alpha diversity comparisons between healthy volunteers (HV), at baseline before bDMARD treatment (B1), and after bDMARDs (B2). **(B)** stool microbiome alpha diversity compared between HV, at baseline before MTX treatment (M1), and after MTX (M2). **(C)** Skin microbiome alpha diversity comparisons between HV, B1, and B2, and **(D)** between HV, M1, and M2. Stool microbiome beta diversity (Bray-Curtis) comparison between **(E)** HV, B1, and B2, and **(G)** HV, M1 and M2. Unweighted UniFrac comparisons between **(F)** HV, B1 and B2, and **(H)** HV, M1 and M2. Skin microbiome beta diversity (Bray-Curtis) (**I**,**K)** and Unweighted UniFrac, comparisons **(J**,**L)** with arrangement similar to stool microbiome. Healthy volunteer data used as a control group in both M1/M2 and B1/B2 comparisons. Coloured areas represent standard errors of the group ordination values. Lines connect two categories where the differences were significant with * (P < 0.05), ** (P < 0.01), or *** (p < 0.001).

When comparing alpha diversity of stool samples from PsA individuals before and after MTX, or bDMARD treatment, neither the Shannon nor the richness measures revealed any differences in the overall diversity or evenness of the stool microbiota after treatment (M1/M2 and B1/B2).

In contrast, while a reduction in the alpha diversity was observed in the skin microbiota before and after treatment in PsA samples this was not statistically significant. However, the reductions in both the Shannon index and richness observed in the post-treatment skin samples (3-6 months after treatment with MTX (M2) and bDMARD groups (B2) when compared to healthy control samples was significant (Figure 1:C-D).

The beta diversity, a comparative measure of the diversity of microbial species between groups, were marginally significant for the stool samples, being only observed for the Unweighted UniFrac in those individuals being treated with biologics. (Figure 1: E-H). On the other hand, the differences in beta diversity measures for skin samples were significant and consistent for the Bray-Curtis and Unweighted UniFrac indices between groups in both MTX and bDMARD cohorts (∼5% in HV/M1/M2 and ∼4% in HV/B1/B2). PERMANOVA analysis also revealed significant but marginal expected differences in the beta diversity between dry and sebaceous skin sites that accounted for ≤ 1% of the variation. (Figure1: I-L).

### Differences in bacterial populations between stool sample groups

In the stool samples, consistent differences were observed in bacterial populations, identified as Operational Taxonomic Units (OTUs), in PsA before and after treatment (MTX or bDMARD) when compared to the healthy controls. Differences were seen when using all the OTU data (partial least squares discriminant analysis PLS-DA) and when analysis was restricted only to discriminant OTUs (sparse PLS-DA -sPLS-DA). Specifically, there were no differentially expressed OTUs between PsA stool samples before and after treatment with bDMARD (Figure 2A-B), but both these PsA groups have OTUs that enable them to be differentiated from the stool microbiota of healthy volunteers (Figure 2C). Despite no statistically significant differences in the diversity of stool samples from PsA donors before and after treatment with MTX, several OTUs were enriched, and both these groups are consistently different from healthy controls (Figure 2:D-F).

**Figure 2:**
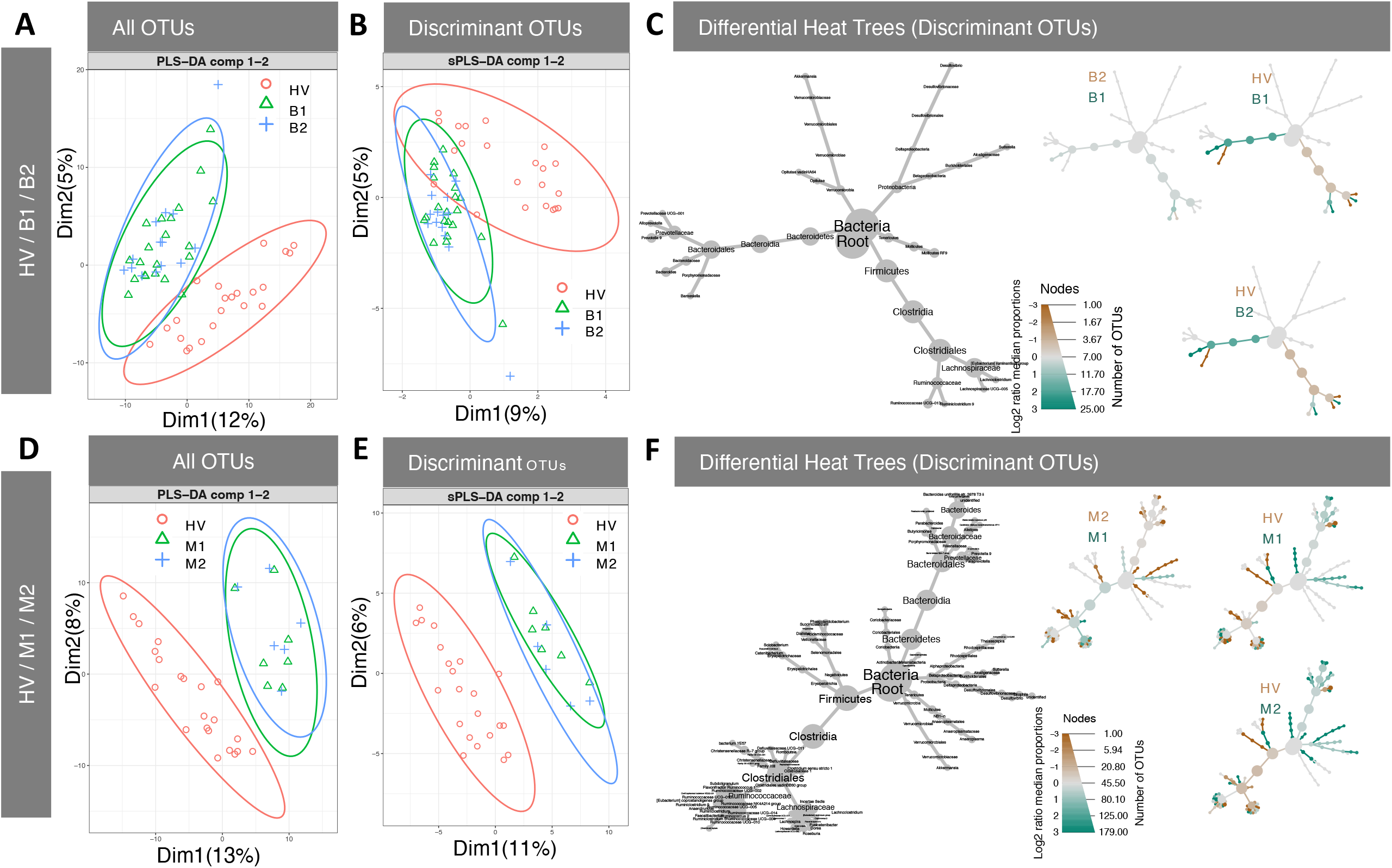
Identification of discriminant OTUs in stool samples. Partial least squares-discriminant analysis (PLS-DA) and sparse PLS-DA (sPLS-DA) of stool samples reveal differences in OTUs between PsA and HV samples **(A)** PLS-DA and **(B)** sPLS-A of HV vs B1 and B2 stool samples. **(C)** shows the differential heat tree identifying the changed OTUs between HV, B1, and B2 samples, with colour-coding on the individual comparison trees showing direction and magnitude of changes in pairwise comparisons. **(D)** PLS-DA and **(E)** sPLS-A of HV vs M1 and M2 stool samples. **(F)** shows differential heat trees for HV, M1 and M2 samples. The same healthy volunteer data is used as a control group in both M1/M2 and B1/B2 comparisons.

Differential heat trees generated for visualisation revealed that both pre and post treatment with bDMARDs the stool samples in the PsA cohort are enriched for *Bacteriodes*, Firmicutes *Lachnoclostridium* and *Ruminiclostridium 9* and depleted in *Barnesiella, Lachnospiraceae UCG-005* and Eubacteria (*ruminantium* group) compared to healthy controls (Figure 2C). In the MTX PsA cohort (pre and post treatment) comparisons with healthy controls reveal a more complex pattern. As with the bDMARD samples, the pre and post MTX samples are enriched in *Bacteriodes genera* but depleted in *Lachnospiraceae* and *Alistipes*. In addition, bacteria in the Clostridiales order including those belonging to the family Christensenellaceae and Ruminococcaceae were enriched after treatment with MTX (M2 vs M1); these Clostridiales were also enriched in the HV group compared to both the pre- and post-treatment MTX cohorts. In contrast to the bDMARD PsA group, the MTX group was enriched in *Proteobacteria* when compared to healthy controls (Figure 2F). Thus, despite the fact that assessments of alpha and beta diversity measures do not reveal significant differences in the stool samples in PsA as a result of treatment, consistent changes in specific bacterial populations were observed when PsA groups were compared to HV samples. Differences between HV and PsA samples were more prominent in the MTX cohort than in the bDMARD cohort.

### Changes in bacterial populations on psoriatic skin plaques after bDMARD or MTX treatment

The ordination plots (PLS-DA and sPLS-DA) show consistent differences between healthy controls and the microbiota on the pre-treatment psoriatic plaques of people with PsA. However, unlike the stool samples, the skin OTUs were observed to change after treatment with both bDMARDs (Figure 3:A-B) and with MTX (Figure 3:C-D). Differential heat trees Figures 3:E-F show the discriminant OTUs identified within each skin site (sebaceous or dry) for each of the three pairwise comparisons between healthy controls and pre and post treatment groups. This analysis reveals some of the complex and consistent differences in the communities of skin bacteria found on the skin surface of healthy controls and PsA samples.

**Figure 3:**
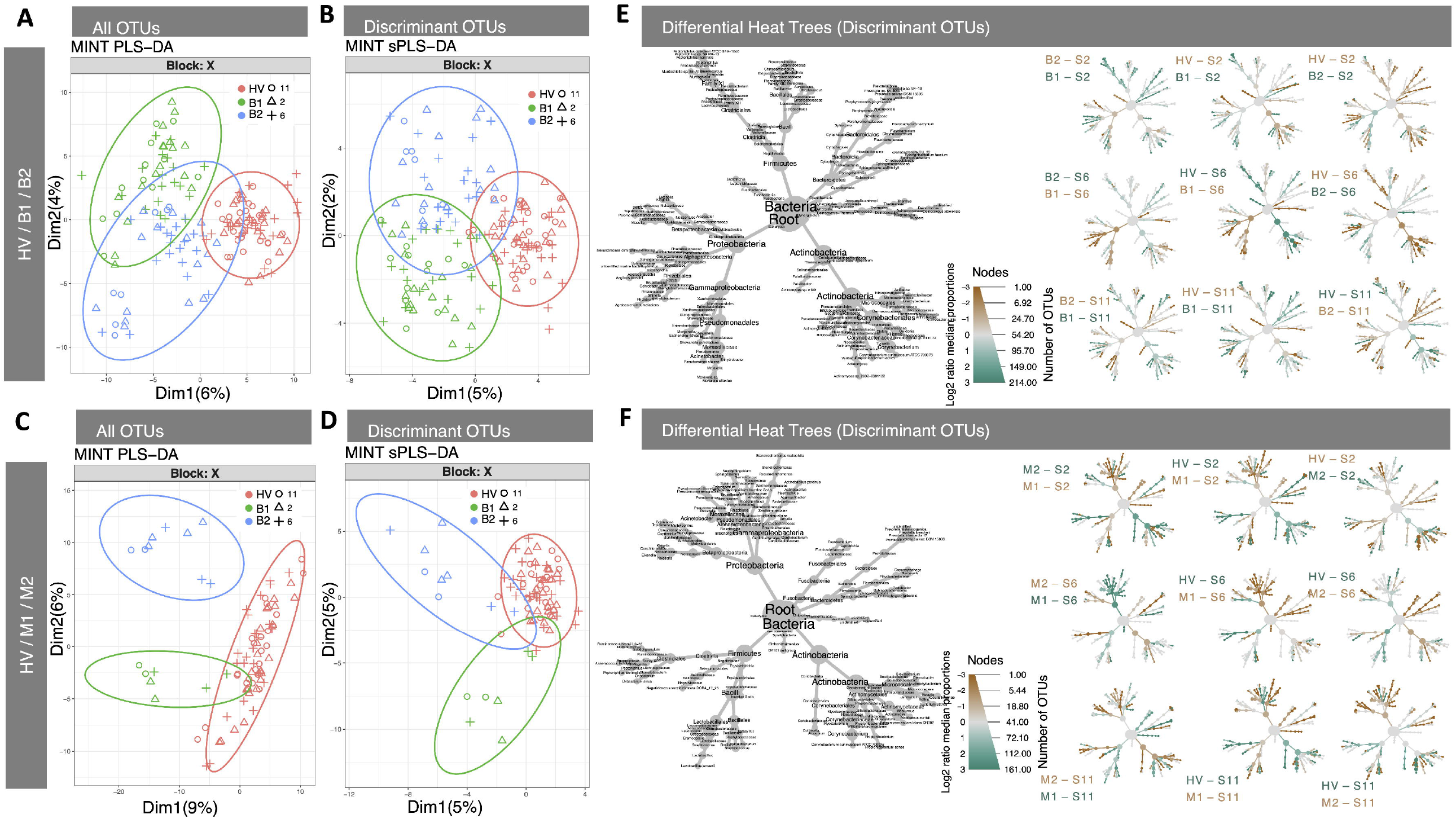
Identification of discriminant OTUs in samples from skin. Partial least squares-discriminant analysis (PLS-DA) and sparse PLS-DA (sPLS-DA) of skin swab samples analysed using MINT reveal differences in OTUs between PsA and HV samples **(A)** PLS-DA and **(B)** sPLS-A of HV vs B1 and B2 skin samples. **(C)** PLS-DA and **(D)** sPLS-A of HV vs M1 and M2 skin samples. **(E)** shows the differential heat tree identifying the changed OTUs between HV, B1, and B2 samples, with colour-coding on the individual comparison trees showing direction and magnitude of changes in pairwise comparisons. **(F)** shows differential heat trees for HV, M1 and M2 samples. The same healthy volunteer data is used as a control group in both M1/M2 and B1/B2 comparisons. Skin sampled at three sites; two were dry (S2, S11) and one sebaceous (S6).

OTUs that were observed to discriminate between all skin sites from individuals with PsA pre-treatment and healthy individuals and post-treatment PsA were *Capnocytophaga, Exiguobacterium, Veillonellaceae, Cardiobacterium, Stenotrophomonas*. Clades identified as being less abundant in individuals with PsA before treatment, compared to healthy skin microbiota, included *Micrococcales*, and *Corynebacterium*. Despite the relatively small numbers of samples in the MTX cohort, the discriminant analysis reveals that there is a depletion of Corynebacterium at all three skin sites when compared to healthy individuals before treatment, which remains lower in the post MTX treatment cohort when compared to healthy individuals. However, in the dry skin microenvironment after MTX treatment there were a few (*Alloscardovia, Mycobacterium* and *Brachybacterium*) clades of the Actinobacteria phylum in higher abundance in PsA individuals when compared to healthy volunteers. After treatment with MTX the taxon *Micrococcales* appeared more abundant in the dry skin microenvironment, similar to what is observed in the healthy skin. However, in the dry skin microenvironment *Alloscardovia* showed increased in abundance when compared to healthy controls. (Figure 3F).

Comparison of bacteria between healthy skin and PsA participants pre-bDMARD treatment revealed that several OTUs belonging to the *Acinetobacter* taxa were observed to discriminate between groups, all being increased in the PsA cohort samples at all sampled sites. Conversely, several OTUs from the Actinobacteria phylum identified as *Actinobacteria, Corynebacterium, Corynebacteriaceae, Dermacoccus* and *Propionibactreium acne* and *Flavobacterium* (Bacteroidetes) and *Anaerococcus* (Firmicutes) were decreased in the pre-bDMARD samples. When comparing skin PsA samples before and after bDMARD treatment, the previously increased clades *Acinetobacter* and *Actinomyces* fell towards levels observed in healthy controls. While no increases were observed after bDMARD treatment in any of the clades observed to be reduced pre-treatment, the Micrococcaceae taxa decreased in the dry skin micro-environment after bDMARD treatment.

Taken together these analyses demonstrate that, while individuals with PsA do not show consistent differences in the alpha or beta diversity of their stool or skin microbiota, discriminant analysis does identify organisms that are consistently different between healthy volunteers and individuals with PsA before initiation of treatment. As described above, many of the discriminating baseline skin PsA microbiota reverted towards levels observed in healthy volunteers after MTX or bDMARD treatment. However, neither treatment had detectable effects on stool microbial populations in PsA.

### Identifying associations between changes in microbiota and the immune response

We assessed frequencies of circulating T cells, B cells, NK cells, dendritic cell, and monocyte populations by flow cytometry. Individuals with PsA had an altered systemic immunophenotype; baseline PsA bDMARD samples displayed increased frequencies of total leukocytes (live CD45^+^), T cells (CD4^+^ and CD8^+^), B cells (CD19^+^), and monocyte populations (MHC II^+^, CD14^-^ CD16^-^ / CD14^+^ CD16^-^ / CD14^+^ CD16^+^) (Figure 4A-C).

**Figure 4:**
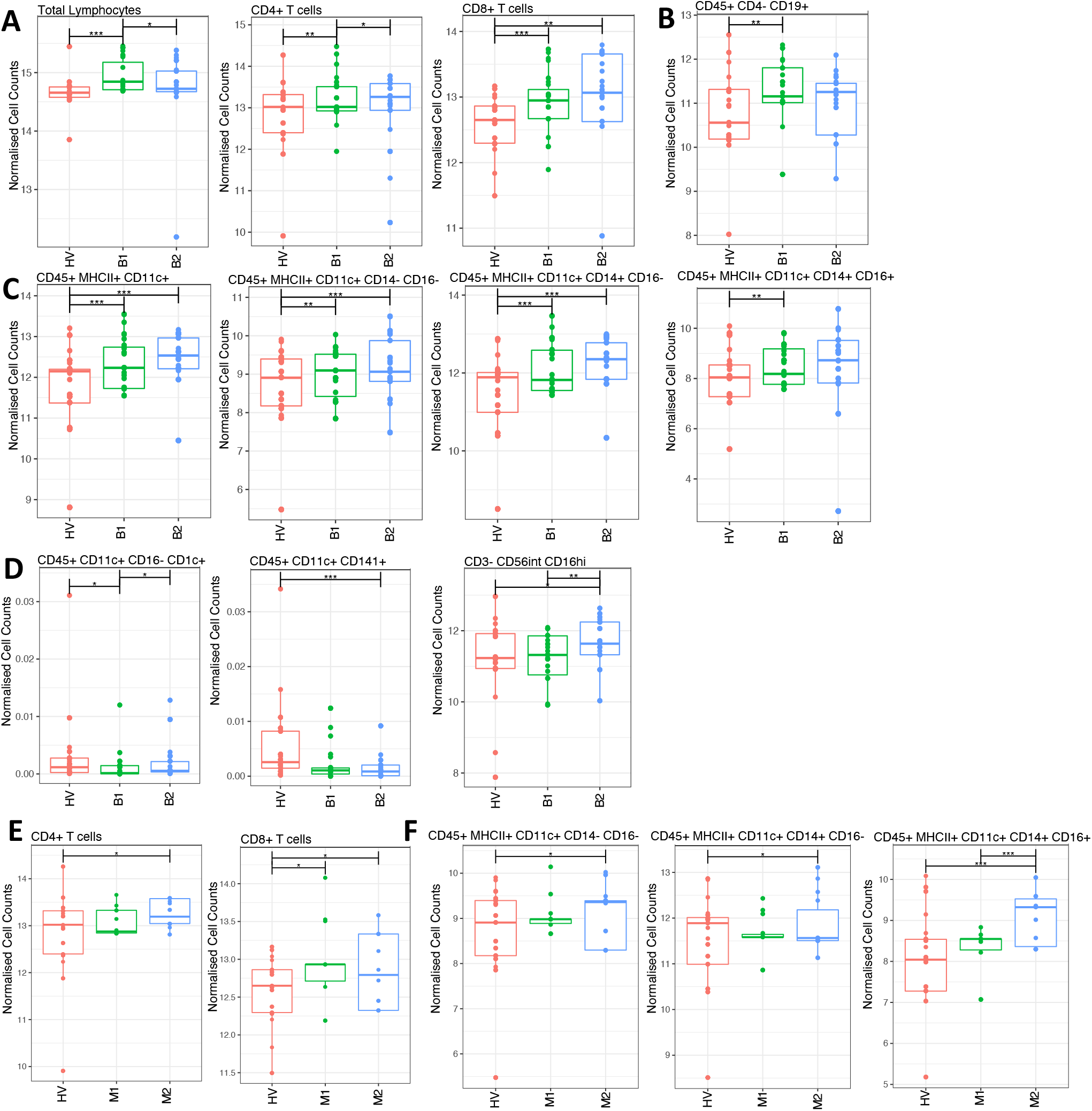
Analysis of peripheral immunophenotype reveals disease- and treatment-associated changes. Blood samples were collected and analysed using flow cytometry to assess the frequencies of circulating immune cells. Absolute cells numbers were assessed, data were normalised, and comparisons made between HV and bDMARD-treated PsA patients **(A-D)** between HV and MTX-treated PsA patients **(E-F)**. Populations with significant changes are shown. These included T cells **(A, E)**, B cells **(B)**, monocytes **(C, F)**, dendritic cells and NK cells **(D)**.

Conversely, frequencies of populations of both circulating conventional dendritic cells and cytotoxic NK cells were lower in PsA patients than in the HV group (Figure 4D). In addition, as expected, after treatment with bDMARDs the frequencies of total lymphocytes and B cells reverted towards the HV baseline values, while frequencies of CD4+ and CD8+ T cells, monocytes, and NK cells remained increased (Fig. 4A-C). In PsA patients treated with MTX, where fewer samples were available and disease activity was milder, fewer differences in the immunophenotype were observed (Figure 4E-F). Consistent with the bDMARD groups, individuals with PsA treated with MTX displayed higher frequencies of CD4 and CD8+ T cells, and of monocyte populations, before and after treatment.

The first step in identifying any associations between the changes in the microbiome and the immune response was to identify the discriminant features that differ when all groups are compared together (shown in Figure 5A for stool). These were compared for correlations between discriminant OTUs and discriminant features of the immunophenotype (Figures 5B), as shown on the differential heat trees. DIABLO analysis revealed that the increased frequency of *Bacteriodes* in stool samples from participants with PsA were significantly and positively correlated with changes in both CD4+ and CD11c+CD14+CD16-PBMC populations (Figure 5B, C) which is sustained even after bDMARD treatment. Furthermore, the DIABLO analyses revealed the changes in the frequencies of the Clostridiales family/clade, specifically Lachnoclostridium and Ruminiclostridium and of other OTUs identified as Clostridiales, enriched in the stool samples from the bDMARD cohort (pre and post treatment) are also associated with alterations in the PBMC immunophenotype. These microbial changes persist after treatment and are significantly associated with changes in patients’ PBMC immunophenotype, which also persist after treatment.

**Figure 5:**
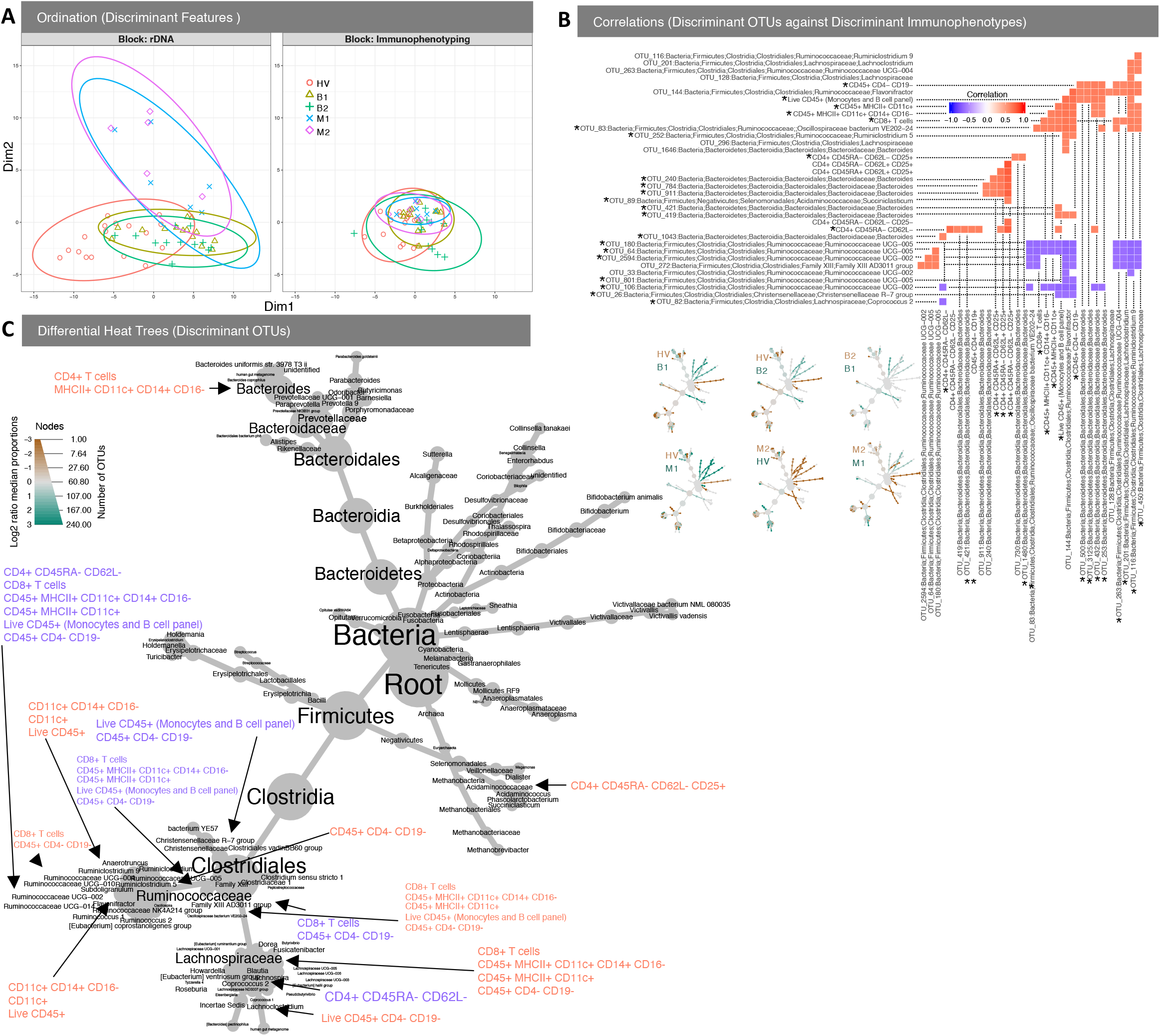
DIABLO analysis comparing changes in stool microbiota and immunophenotype reveals significant associations. **(A)** The algorithm found two components (minimum number of components to find OTUs that discriminate between all conditions) reducing the classification error rates in the DIABLO algorithm (using Centroid distance) and shows the reduced order representation of samples with ellipses representing 95% confidence interval and percentage variations explained by these components in axes labels for both microbiome (Block: rDNA) and immunophenotyping (Block: Immunophenotyping). **(B)** Correlations whether positive (red) or negative (blue) between microbiome and immunophenotype returned from the algorithm. Annotation of “star” is used only when the relationship is between microbiome and immunophenotype and not otherwise. **(C)** shows where on the resultant tree are microbes affected by immune cells with the colors same as shown in (B).

A similar analysis of the skin microbiota and the PBMC immunophenotype (Figure 6A) found a number of associations between the frequency of CD8+ CD45RA-CD62L-(Effector memory) T cells and OTUs among the Actinobacteriaceae (Figure 6B). Many of these OTUs are observed at higher frequencies in PsA samples before treatment and fall after treatment, as described previously (Figure 3). However, the CD8+ T cell frequency remains high in PsA after treatment. Thus, the observed association between Actinobacteriaceae and CD8+ T cells relates to the differences observed between HV and PsA pre-treatment samples.

**Figure 6:**
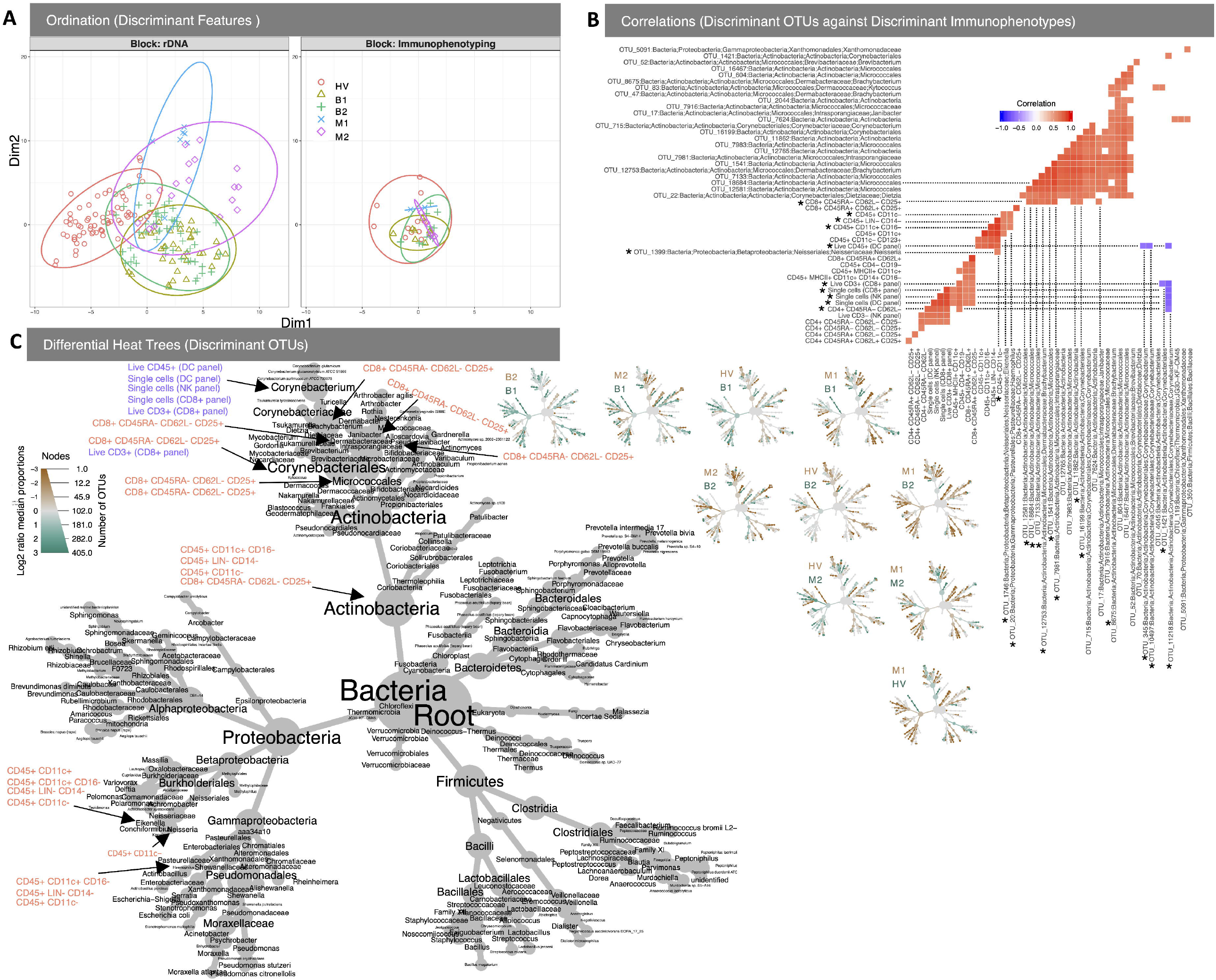
DIABLO analysis comparing changes in skin microbiota and immunophenotype reveals significant associations. **(A)** The algorithm found five component (minimum number of components to find OTUs that discriminate between all conditions) reducing the classification error rates in the DIABLO algorithm (using Centroid distance) and shows the reduced order representation of samples with ellipses representing 95% confidence interval and percentage variations explained by these components in axes labels for both microbiome (Block: rDNA) and immunophenotyping (Block: Immunophenotyping). **(B)** Correlations whether positive (red) or negative (blue) between microbiome and immunophenotype returned from the algorithm. Annotation of “star” is used only when the relationship is between microbiome and immunophenotype and not otherwise. **(C)** shows where on the resultant tree are microbes affected by immune cells with the colors same as shown in (B).

## Discussion

This is the first prospective study into longitudinal associations of stool and skin microbiota with treatment in PsA and the relationship with immunophenotype; we explored associations between skin and stool bacteria and the immunophenotype of PsA patients, before and after treatment with MTX or bDMARDs. Healthy controls were included as comparators. We find that while alpha and beta diversity of stool and skin microbiota in PsA before treatment are not significantly different from healthy controls, specific OTUs are consistently different. Treatment with either therapy affects the skin but not the stool microbiota, revealing persistent abnormalities in stool microbiota in PsA. Patients with PsA also have an altered systemic immunophenotype, which, whilst altered by treatment, does not revert to normal. Finally, significant correlations between bacterial taxa in individuals’ stool and their systemic immune parameters reveal connections between persistent underlying intestinal changes and systemic immune responses that are not affected by therapy.

Using sPLS-DA analysis, we identified stool microbial features that discriminated between PsA groups and healthy samples, including *Bacteroides*, which was the most abundant taxon in the PsA patients. This finding is consistent with previous studies, which have reported that *Bacteroides* also changes with diet, lifestyle (13) and disease state including in enthesitis-related arthritis(14). In stool, apart from *Bacteroides*, the discriminant OTUs were largely from low abundance taxa. Our findings are consistent with previous studies. For example, *Bacteroides* the most abundant taxon in the gut of our PsA patients, also changes with diet, lifestyle(13) and disease state including in enthesitis-related arthritis(14). The discriminatory low abundance taxa in the bDMARD group may merit further exploration, as some have been identified in other disease states. *Proteobacteria, Barnesiella* and *Lachnospiraceae*, for instance, are potential biomarkers for treatment responses(15-17) or pathology (18).Interestingly, *Lachnoclostridium, Ruminiclostridium*_9 and *Bacteroides* were down-regulated in stool samples from the PsA bDMARD cohorts compared to healthy controls. These organisms have been associated with modulation of lipid metabolism, reduced inflammation, and improved intestinal barrier function in mice treated with reserveratol(19). The discriminating taxa that increased in our MTX PsA groups contained butyrate producers (*Eubacteria, Ruminococcus, Christensenellaceae, Alistipes*) that have been reported to facilitate positive immunomodulatory effects (20, 21)

The skin microbiota has not been extensively studied in PsA. In psoriasis observations have been mixed(21), and “cutaneo-types”(3) dominated by major phyla have been reported. We did not find significant differences in microbial alpha diversity between PsA samples and healthy controls. However, discriminatory, low abundance, skin taxa were identified that change post treatment; these contained *Actinobacteria*-associated taxa. Interestingly, the *Acinetobacter* that increased in PsA lesional skin prior to bDMARD treatment are reported to associate with expression of anti-inflammatory genes in healthy skin(22). Therefore, the association of these taxa with psoriatic skin may be of interest.

Treatment with bDMARDs affected the systemic immunophenotype, with a return to normal levels for total leukocyte numbers. However, many immune cells were found at higher frequencies even after treatment. Fewer MTX-treated individuals were analysed, and changes appear less pronounced in this group. However, after MTX treatment there were also increases in the frequency of CD4^+^ T cells and in populations of circulating monocytes. Thus, even after treatments, systemic changes in the immunophenotype persisted, consistent with continuing systemic inflammation.

We explored variations between individuals to identify interactions between the immunophenotype and microbiota and found an association between *Bacteroides* and CD4^+^ T cells, consistent with previous reports(23). We also found a correlation between CD11c^+^ cells (dendritic cells) and *Bacteroides* previously observed in Crohn’s disease(24), and novel correlations with *Ruminoclostridium* and *Lachnospiraceae*. In PsA, therefore, changes occur in stool bacterial taxa. These persist after treatment and are associated with specific changes in circulating T cell and myeloid cell populations. In the skin, increases in *Actinobacteria*, especially *Corynebacterium*, correlated with increased frequencies of circulating CD8 cells. *Corynebacteria* have been shown to drive immune responses in mouse skin(25). Thus, there may also be an association in humans between specific lesional bacteria and circulating immune cells. Limitations of the study include the modest sample size, and the follow-up of only 3-6 months. A longer follow-up could identify additional changes in the gut microbiota that lag further behind the treatment.

In summary, we find that PsA-associated changes in stool microbiota were not affected by MTX or bDMARD treatment. In contrast, a marked effect of treatment was observed on the skin microbiota. Further work investigating the correlations between persistent dysbiotic stool microbiota and the immunophenotype we have identified may reveal potential microbial targets to enhance treatment responses through adjunctive therapies in PsA.

## Data Availability

All data produced in the present study are available upon reasonable request to the authors

## Acknowledgements and affiliations

This work was funded by the Versus Arthritis Microbiome Pathfinder Award (grant ref 21130) and by the NIHR Manchester Biomedical Research Centre; the views expressed in this publication are those of the author(s) and not necessarily of the NHS; the National Institute for Health research or the Department of Health. Thanks to Diane Vaughan at the Flow Core Facility at the Institute of Infection, Immunity and Inflammation. AB is an NIHR Senior Investigator.

