## Supplemental information for "The stool microbiome in patients with psoriatic arthritis is altered but, unlike the skin microbiome, does not change following treatment: evidence for an underlying inflammatory drive from the intestine"

**Supplementary Information**

### Statistical Methods

Statistical analyses were performed in R using the combined data generated from the bioinformatics as well as meta data associated with the study. The vegan package [1] was used for alpha (diversity() function) and beta diversity analyses. Ordination of OTU table in reduced space (beta diversity) was done using Principal Coordinate Analysis (PCoA) plots of OTUs using two different distance measures were made using Vegan's cmdscale() function: (1) Bray-Curtis is a distance metric which considers only OTU abundance counts, and (2) Unweighted Unifrac is a phylogenetic distance metric which calculates the distance between samples by taking the proportion of the sum of unshared branch lengths in the sum of all the branch lengths of the phylogenetic tree for the OTUs observed in two samples, and without taking into account their abundances. Unifrac distance was calculated using the phyloseq package [2].

Analysis of variance (referred to as PERMANOVA) for explanatory variables (or sources of variation) was performed using Vegan's adonis() against distance matrices (Bray-Curtis/UnweightedUniFrac). Discriminant analyses were performed using three different algorithms. For the stool samples, we used Sparse Projection to Latent Structure – Discriminant Analysis (sPLS-DA) from the R's mixOmics package [3]. The procedure constructs artificial latent components of the predicted dataset (denoted as $X(N \times P)$) and the response variable (denoted as $Y$ m with categorical information of samples, e.g. treatment response on methotrexate, HV1, M1, M2, and treatment response on biologics, HV 1, B1, B2) by factorizing these matrices into scores and loading vectors in a new space such that the covariance between the scores of these two matrices$\mathrm{cov}(X_h a_h, Y_h b_h)$ in this space is maximized under two constraints: $\|a_h\|_2 = 1$; and $\|a_h\|_1 \leq \lambda$, where $a_h$ and $b_h$are the corresponding loading vectors for $X$ and $Y$, and $h$ represents the number of components (akin to PCA analysis). The first constraint ensures the loading vector to have unit magnitude (requirement of the procedure) and the second constraint (also called $l_1$ penalty) to ensure that for the features that do not vary between the categories, the corresponding loading vector coefficients go to zero. This is done by using the sparsity control parameter $\lambda$ in the above equation, and by adjusting it enforces shrinkage of loading vector coefficients. According to the recommendations given in mixOmics package (http://www.mixomics.org), before applying the procedure splsda(), we pre-filter 1% of the lowest abundant genera and then perform TSS+CLR (Total Sum Scaling followed by Centralised Log Ratio) normalisation. To predict the number of latent components (associated loading vectors) and the number of discriminants, the perf.plsda() and tune.splsda() functions were used, respectively. In the latter case, we fine tune the model was applied using leave-one-out cross-validation by splitting the data into training and testing sets and then finding the classification error rates employing two metrics.

For the skin samples, where samples were collected from three sites (Site:11, Site:2, Site:6), we have used the Multivariate Integration (MINT) algorithm [3], which is an extension of the multi-group Projection to Latent Structure (mgPLS), and it attempts to find a common projection space across all studies, defined on a small subset of discriminative variables that consistently discriminate the Skin sites (Site:11, Site:2, Site:6). In MINT, we have combined $M = 3$ datasets denoted $X^{(1)}(N_1 \times P)$, $X^{(2)}(N_2 \times P)$, $X^{(3)}(N_3 \times P)$ for Site:11, Site:2, Site:6, respectively, where all the datasets share the P OTUs whilst the number of samples differ, i.e., $N_1$, $N_2$, $N_3$. All three sites have associated dummy indicator outcome $Y^{(1)}$, $Y^{(2)}$, $Y^{(3)}$ in which all the time points (, e.g. treatment response on methotrexate, HV1, IMIGPA PsA M1, IMIGPA PsA M2, and treatment response on biologics, HV 1, IMIGPA PsA B1, IMIGPA PsA B2) are represented. MINT then solves the problem: $\max_{a_h, b_h} \sum_{m=1}^{M} N_m \mathrm{cov}(X_h^{(m)} a_h, Y_h^{(m)} b_h)$, with the previous constrains$\|a_h\|_2 = 1$ and $\|a_h\|_1 \leq \lambda$, where the covariance of scores between the datasets are maximised by finding the global loading vectors $a_h$ and $b_h$ common to all studies.

The prefiltering and the cross-validation procedure is similar to the previous case (sPLS-DA). To predict the number of latent components (associated loading vectors) and the number of discriminants, the mint.plsda() and tune () functions were used, respectively.

Discriminant analyses between the microbiome and immunophenotypes was done using DIABLO from R's mixOmics package [3] was used. We have combined $M = 2$ datasets denoted $X^{(1)}(N \times P_1)$, $X^{(2)}(N \times P_2)$ where $X^{(1)}$ represents the microbiome data, and $X^{(2)}$ represents immunophenotyping data. The algorithm then constructs artificial latent components of the datasets by factorizing the datasets into scores and loading vectors in new space such that the covariance between the scores of these matrices in this space is maximized, i.e., for $q = 1,2,\ldots,Q,$ DIABLO solves for each component $h = 1,\ldots,H$:

$$\underset{a_h^{(1)},\ldots,a_h^{(Q)}}{\mathbf{arg\,max}} \sum_{q,j=1,q\neq j}^{Q} c_{q,j} \operatorname{cov}\left(X_h^{(q)} a_h^{(q)}, X_h^{(j)} a_h^{(j)}\right) \quad \text{s.t.} \quad \left\|a_h^{(q)}\right\|_2 = 1 \text{ and } \left\|a_h^{(q)}\right\|_1 \leq \lambda^{(q)}$$

where $\lambda^{(q)}$ is the penalization parameter, $a_h^{(q)}$ is the loading vector on component $h$ associated to the (deflated) matrix $X_h^{(q)}$ of the data set $X^{(q)}$, and $C = \{c_{q,j}\}_{q,j}$ is the design matrix. $C$ is a $Q \times Q$ matrix that specifies whether datasets should be correlated and includes values between zero (datasets are not connected) and one (datasets are fully connected). Discriminant analysis is thus possible by replacing one data matrix $X^{(q)}$ with the outcome dummy matrix $Y$ (indicating the labelling of samples in this study as HV1, IMIGPA PsA B1, IMIGPA PsA B2, IMIGPA PsA M1, IMIGPA PsA M2 ). The constraints $\left\|a_h^{(q)}\right\|_2 = 1$ and $\left\|a_h^{(q)}\right\|_1 \leq \lambda^{(q)}$ have a similar definition as in the case of sPLS-DA. We have used the same prefiltering strategy and cross-validation procedure. For design matrix, mixOmics suggests that a *full weighted design* where $c_{q,j} = 0.1$ between data matrices (microbiome and immunophenotyped) and 1 for the outcome dummy matrix (HV1, IMIGPA PsA B1, IMIGPA PsA B2, IMIGPA PsA M1, IMIGPA PsA M2) leads to a trade-off between maximizing correlation between datasets and maximizing the discrimination of the outcome and therefore we used as such. The prefiltering and the cross-validation procedure is similar to the previous case (sPLS-DA), where as for immunophenotyping data, we have applying arcsinh transformation to flowcytometry cell counts as is the general practise. To predict the number of latent components (associated loading vectors) and the number of discriminants, the block.splsda() and tune.block.splsda() functions were used, respectively.
